# COVID-19 and Influenza: Vaccination Before and During the Pandemic among the Lebanese Adult Population

**DOI:** 10.1101/2021.02.10.21251392

**Authors:** Anthony Elia, Nedal Taha, Sima Tokajian

## Abstract

Influenza is a common respiratory tract disease that has been around for years. Vaccination remains the most cost-effective measure to avoid infection. Influenza vaccination rates in Lebanon, like elsewhere in the world, are known to be suboptimal. The emergence and spread of SARS-CoV-2 led to a global health crisis. This study aimed at assessing the impact of the COVID-19 pandemic on the tendency of the general adult population residing in Lebanon to vaccinate against influenza. A quantitative cross-sectional study was conducted between November and December 2020 using a structured questionnaire passed on 1055 individuals to determine factors influencing vaccination habits prior to and during the COVID-19 pandemic. The factors were analyzed using univariate, bivariate, and multivariate analyses. The majority (69.7%) of the study participants never received the influenza vaccine within the last 5 years, 20% vaccinated occasionally, and only 10.3% were yearly vaccinated. Among individuals who never got the influenza vaccine within the last 5 years, 20.7% reported their willingness to vaccinate this winter significantly increasing the vaccination percentage. Participants concerned about the COVID-19 pandemic showed an enhanced willingness to vaccinate against both. Influenza and COVID-19 vaccination rates, nevertheless, are still considerably lower than the recommended coverage. The COVID-19 pandemic caused a significant increase in the tendency to vaccinate against influenza. Yet, urgent vaccination strategies should be implemented to boost vaccine uptake across all demographics to consequently diminish the burden conflicted by influenza, COVID-19, and ultimately other infectious diseases.

## I. Background

Influenza, a contagious respiratory illness, caused by influenza viruses causes an annual worldwide epidemic resulting in significant morbidity and mortality. The disease is caused by four types of influenza viruses with Influenza A and B being responsible for the seasonal flu epidemics. The disease severity varies from mild upper respiratory symptoms to severe pneumonia and even death (Nafziger and Pratt 2014). Each year, the annual influenza epidemic substantially affects the healthcare causing about 3 to 5 million cases of severe illness, and about 290,000 to 650,000 respiratory deaths (World Health Organization 2018). The degree of genetic shift and drift of the dominant strain as well as the vaccine coverage and efficacy determine the morbidity and mortality.

The emergence and spread of severe acute respiratory syndrome coronavirus 2 (SARS-CoV-2) (Gorbalenya et al. 2020) increased the possibility of facing a “twindemic” of Influenza and Coronavirus Disease 2019 (COVID-19), which could be an unprecedented threat to public health in the wake of globalization.

Although SARS-Cov-2 and Influenza viruses are vastly different pathogens, there is a significant area of overlap. Both predominantly spread via respiratory droplets among people in close contact with non-pharmacologic interventions affecting the incidence of both viruses. However, the management of the two viruses is widely different. Influenza is treated by antivirals such as neuraminidase inhibitors which are not affective against SARS-CoV-2 while remdesivir has been FDA-approved for the treatment of COVID-19 in hospitalized patients (Solomon, Sherman, and Kanjilal 2020). Yet, none of the aforementioned drugs could “eradicate” either one.

Consequently, prevention of these infections through vaccination is the most important cost-effective tool to hinder their spread. The vaccination rate is suboptimal worldwide (Centers for Disease Control and Prevention 2018; European Centre for Disease Prevention and Control 2017). Up till the date of writing the manuscript, the FDA has authorized the Pfizer-BioNtech and the Moderna mRNA COVID-19 vaccine for emergency use (U.S. Food & Drug Administration 2021). Many countries started their vaccination campaigns, with the general population being divided into favorable and unfavorable views on the COVID-19 vaccines (World Economic Forum 2020) largely because of the fast-track witnessed approvals.

Until the 6^th^ of February 2021, we had more than 104 million confirmed cases worldwide with an increased fatality rate in high-risk groups (World Health Organization 2021). The first documented case in Lebanon was on February 21^st^, 2020 and since then we had almost 300,000 confirmed COVID-19 cases and more than 2,500 reported deaths (Ministry of Public Health 2021). Unfortunately, in January 2021, the healthcare system reached its full capacity with complete occupancy in intensive care units. Although all countries worldwide were feeling the strain, Lebanon’s experience was exceptional as this pandemic piled on top of a financial collapse and a huge port blast which already brought the health system to its knees.

There is no national seasonal influenza vaccination program in Lebanon, therefore, the vaccine is paid out-of-pocket. It costs between 8.04$ and 17.41$ (12,115-26,234 Lebanese Pounds) considering the official exchange rate (Ministry of Public Health 2021). The only report evaluating vaccine acceptance in the adult population in Lebanon was that conducted by El Khoury and Salameh (2015). Their results revealed an alarmingly low (27.6%) regular uptake of the influenza vaccine even in the high-risk groups.

The objective of this study was to assess the knowledge, attitudes, and practices of the Lebanese population towards the seasonal influenza and its vaccine, and to evaluate the effect of the COVID-19 pandemic on the influenza vaccination in the general Lebanese adult population. Data generated from this study will help to formulate strategies and campaigns to increase acceptance of the influenza and SARS-CoV-2 vaccines in Lebanon.

## II. Data and Methods

The presented cross-sectional study was revised and approved by the Institutional Review Board (IRB) at the Lebanese American University (approval number LAU.SAS.ST3.4/Nov/2020). Informed consents were obtained electronically from participants after presenting the purpose of the study through a written text prior to the start of the survey questionnaire.

### 1. Study design

A 30-item structured, self-established questionnaire was designed and developed in English and Arabic languages (see appendix 1). The questionnaire included 4 sections: **(1)** general information, **(2)** influenza and influenza vaccine, **(3)** COVID-19, and **(4)** influenza–COVID-19 intersection.

Preceding data collection, a pilot study was carried out on 12 volunteers to evaluate the clarity of the questionnaire after which minor modifications were introduced according to their collective feedback.

### 2. Data collection

The survey was disseminated from November 2020 to December 2020 as an electronic link to a Google form entitled “FLUVACO” using social media applications, namely WhatsApp, Instagram, and Facebook. A total of 1055 participants from all adult age groups and residing in all 8 governorates of Lebanon gave their consent to participate.

### 3. Data Analysis

Statistical analysis was performed using Statistical Package for Social Sciences (SPSS), version 26.0 (IBM Corp, Armonk, NY, USA). The data was analyzed using univariate (descriptive), bivariate (Pearson chi-square test for association and McNemar’s test) as well as multivariate (logistic regression) techniques. p-values in the figures are represented as follows: * (p≤0.05), ** (p≤0.01), *** (p≤0.001). Confidence intervals mentioned in the text are at the 95% level.

## III. Findings

### 1. Sample description

The sample included 1055 participants with 66.2% being females. The most represented age group was 25-34 years old (30.2%), which was in line with the prevalent age group in the Lebanese population (Central Administration of Statistics 2019), while the older age group (45 and above) was among the least represented (15.9%). The geographical distribution over the 8 governorates showed that the majority (49.3%) of the participants resided in Mount Lebanon followed by the capital Beirut (14.6%), Northern Lebanon (10.9%), Southern Lebanon (7.4%), Nabatiyeh (5.1%), Akkar (4.4%), Baalbeck-Hermel (4.2%), and Beqaa (4.0%). The highest percentage (74.2%) was for those having a university degree, and 24.2% of the participants were in health-related fields. Regarding occupation, 55.8% were employed, 19.2% were unemployed, 20.8% were university students, and the rest had other occupations (school students, volunteers or retired). The highest response (63%) came from participants with a low financial standing. Financial standing is based on the household total income partitioned equally among the household individuals (see note 1). The vast majority (79.1%) implicated in their responses a decrease in the purchasing power following the economic crisis in Lebanon (in 2019). The studied population mainly (37.8%) belonged to families of 5 members or more. The majority (79%) consulted physicians only when needed. Concerning vaccines in general, a notable 5.1% considered themselves anti-vaxxers while the majority (51.3%) indicated that it would be dependent on the vaccine and/or production company and the disease.

### 2. Influenza vaccination before and during the COVID-19 pandemic

Prior to the COVID-19 pandemic, 718 (69.7%, CI=66.8–72.5%) of the study participants never received the influenza vaccine within the last 5 years. The remaining 312 (30.3%, CI=27.5– 33.2%) were vaccinated at least once: 206 (20.0%, CI=17.6–22.6%) vaccinated occasionally and only 106 (10.3%, CI=8.5–12.3%) were yearly vaccinated. Regarding winter 2020-2021, 696 (66.4%, CI=63.5–69.3%) implied that they will probably or surely not vaccinate, whereas 352 (33.6%, CI=30.7–36.5%) will probably or surely vaccinate. Interestingly, 20.7% of those who never vaccinated reported their willingness to vaccinate this winter. An exact McNemar test showed a significant increase in the percentage of participants intending to vaccinate this winter, p<0.001.

### 3. General associations with influenza vaccination before and during the COVID-19 pandemic

According to our results presented in figure 1, in the last five years, vaccination was significantly influenced by age, occupation, education level, the field of study, frequency of medical visits, and attitudes towards vaccination.

**Figure 1.**
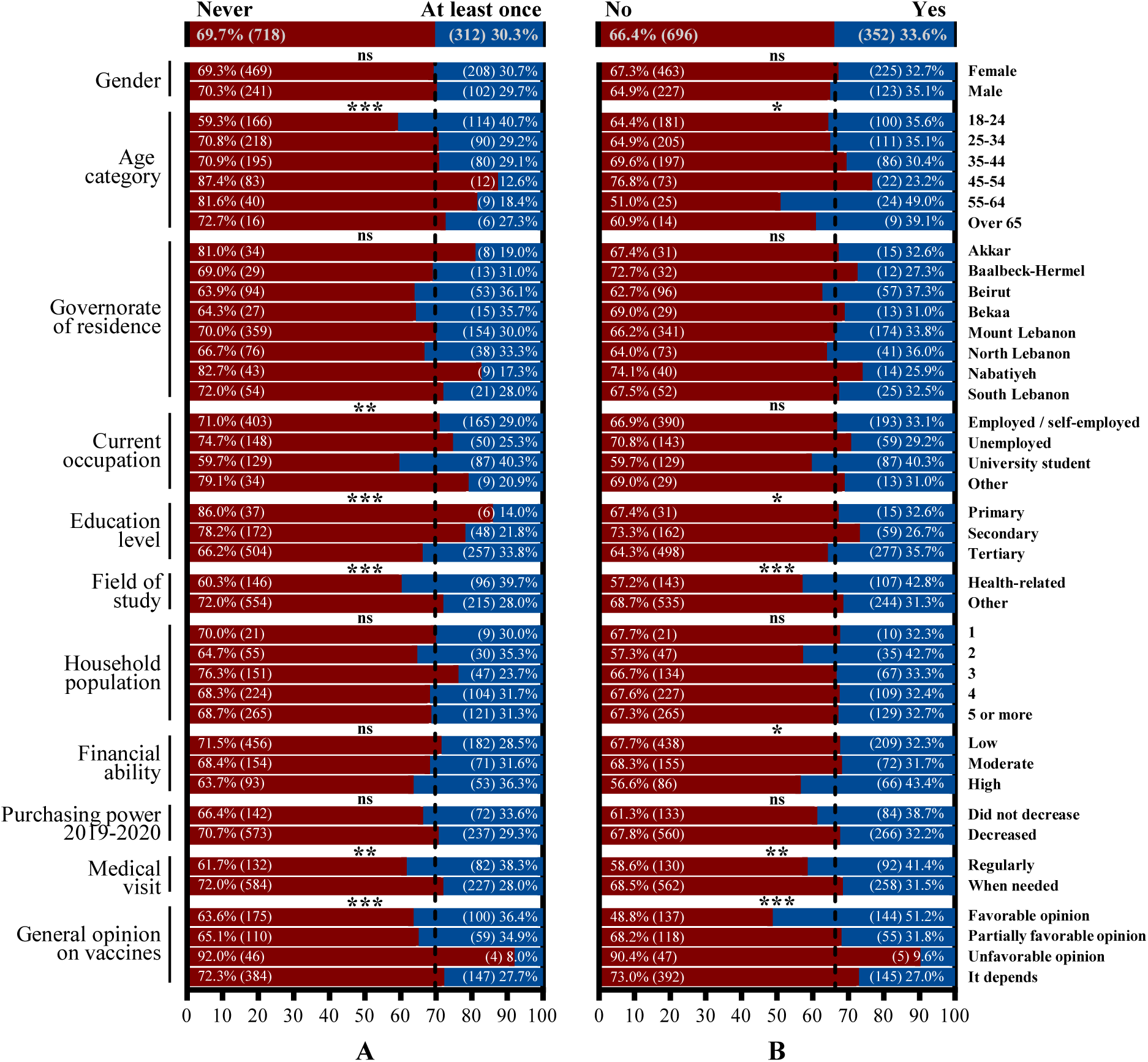
General associations with influenza vaccination (A) within the last 5 years and (B) during the COVID-19 pandemic.

As for the potential to get the influenza vaccine this winter 2020-2021, it was significantly correlated with age, education level, field of study, financial standing, frequency of medical visits, and general opinion on vaccines. Gender, governorate of residency, and household population; however, did not significantly affect participants’ willingness to get the influenza vaccine.

### 4. Knowledge, attitude, and practices regarding influenza and influenza vaccination before and during the COVID-19 pandemic

Figure 2 displays the associations between knowledge, attitude, and practices (KAP) of participants and influenza vaccination **(A)** before COVID-19 and **(B)** during COVID-19.

**Figure 2.**
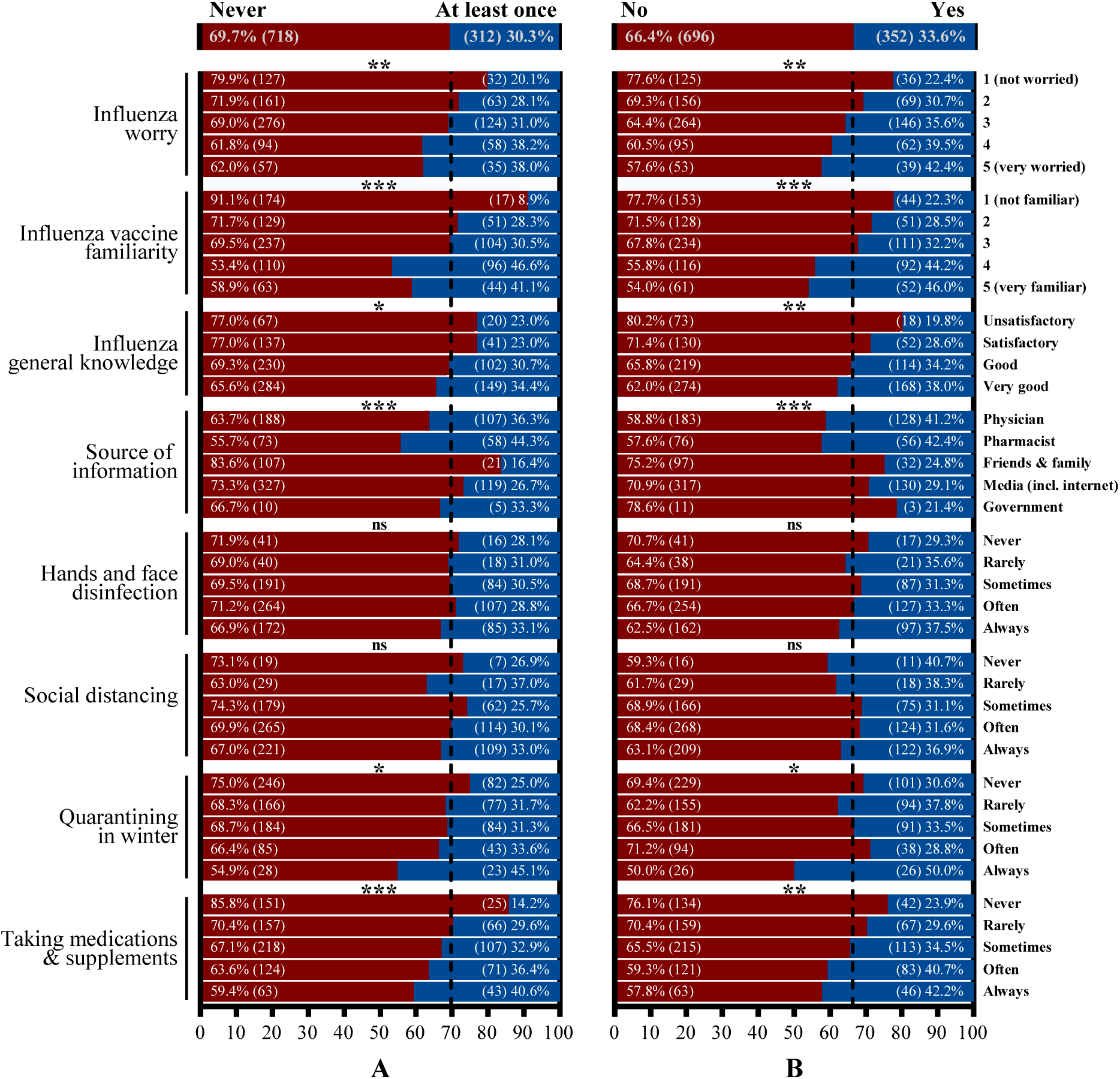
Knowledge, attitude, and practices regarding influenza and influenza vaccination (A) within the last 5 years and (B) during the COVID-19 pandemic.

All the assessed KAP variables have the same relation with influenza vaccination before and during COVID-19. For instance, the more the participants were worried about an influenza infection, the more the participants were familiar with the influenza vaccine (subjective evaluation), and the more general knowledge the participants knew about influenza (objective assessment, see note 2), the significantly higher the percentage of participants vaccinating against influenza.

### 5. Factors associated with influenza vaccination before and during COVID-19 using a multivariate analysis

Figure 3 summarizes the results of the multivariate analysis performed to ascertain the effects of the variables on the likelihood of influenza vaccination. Only the variables that can be targeted were included in the analysis.

**Figure 3.**
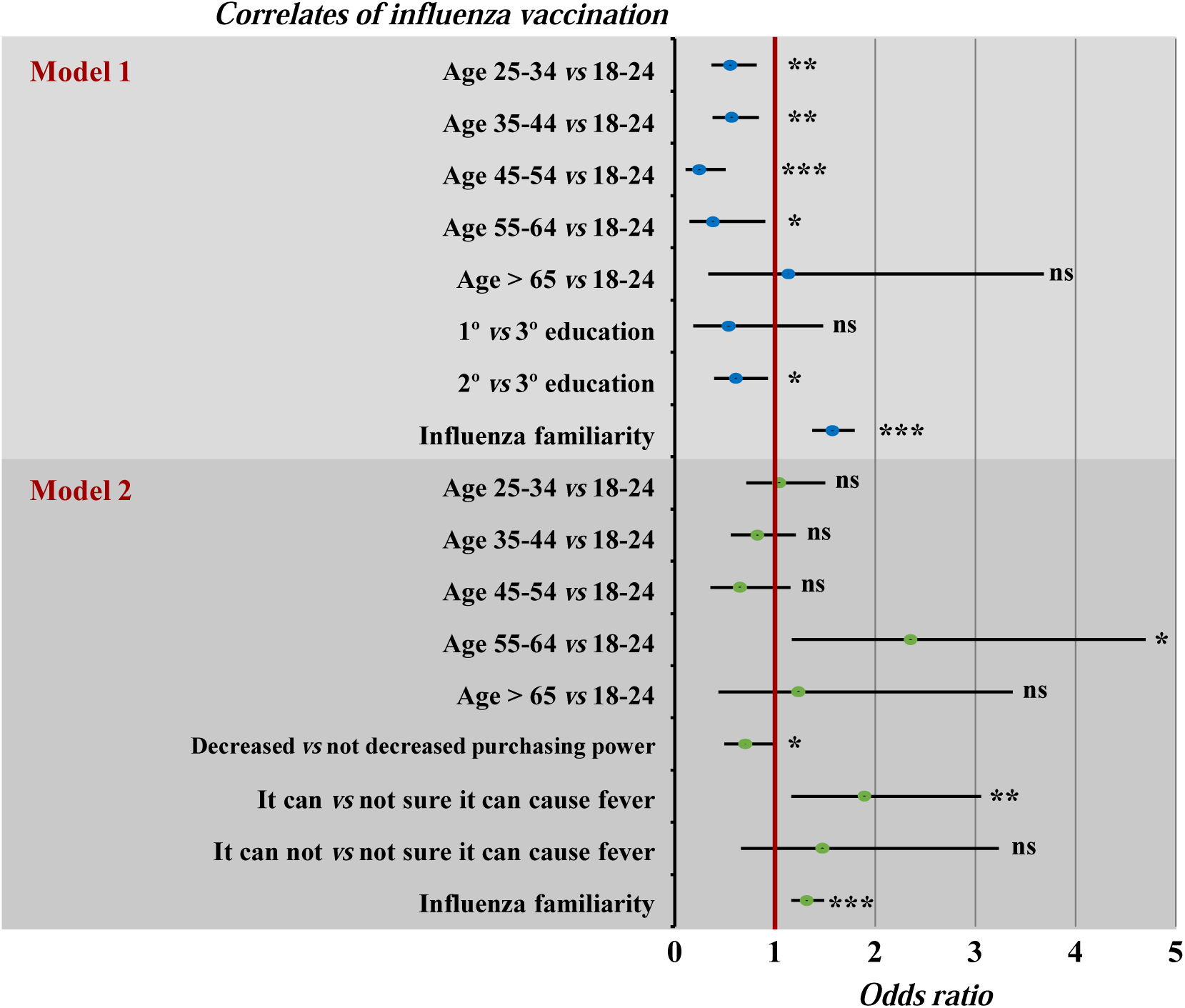
Multivariate analysis of influenza vaccination withing the last 5 years (Model 1) and during (Model 2) the COVID-19 pandemic. Model 1 and 2: In the stepwise forward (WALD) logistic regression to predict influenza vaccination, variables entered were: gender, age, governorate of residency, education level, education background, financial standing, change in purchasing power, and general knowledge. Both logistic regressions were statistically significant (p < 0.001) and explained 7.3% and 8.3% (Nagelkerke R^2^) of the variance in influenza vaccination, respectively.

Model 1 shows that participants in all age categories except above 65 were significantly less vaccinated compared to those aged 18-24. Furthermore, individuals with a secondary education had a lower vaccine uptake relative to those with tertiary education (OR=0.61, CI=0.41–0.91, p=0.016). Also, an increased influenza vaccine familiarity was associated with a significant increase in vaccination rate (OR=1.57, CI=1.38–1.78, p < 0.001).

Model 2 generated for the predicted rate of influenza vaccination this winter 2020-2021 indicates that participants aged 55-64 *vs* 18-24 (OR=2.35, CI=1.18–4.69, p=0.015), who answered correctly that influenza can cause fever *vs* not sure (OR=1.89, CI=1.18–3.04, p=0.008), and are more familiar with the influenza vaccine (OR=1.32, CI=1.17–1.47, p< 0.001) intend to vaccinate more than others this winter. Contrariwise, participants living in households where the purchasing power decreased between 2019 and 2020 are 30% less willing to vaccinate (CI=0.51–0.98, p=0.036) when compared with those living in households where the purchasing power did not decrease.

### 6. COVID-19 effect on influenza vaccination this winter

Participants who were more likely to vaccinate this winter were also more likely to agree that the influenza vaccine should be obligatory, free of charge, and available to everyone (supplementary Figure 1).

Models 1, 2, and 3 presented in Figure 4 were created to assess the effect of the COVID-19 pandemic on the willingness of participants who never, occasionally, or yearly vaccinated, respectively, to get the influenza shot this winter.

**Figure 4.**
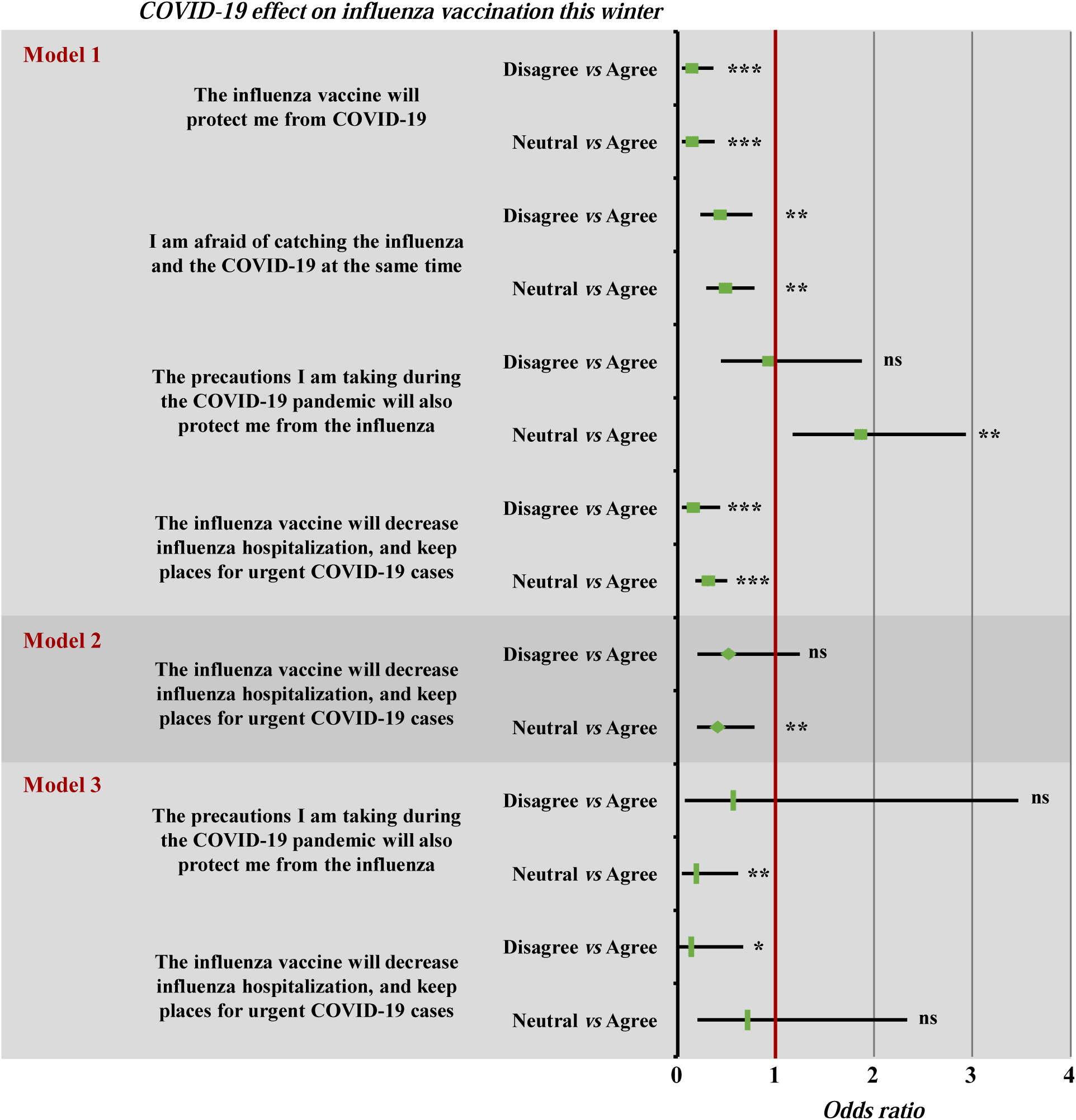
Multivariate analysis: COVID-19 effect on influenza vaccination this winter for those who never (Model 1), occasionally (Model 2), and yearly (Model 3) vaccinated within the last 5 years. Models 1, 2, and 3: In the stepwise forward (WALD) logistic regression to assess the effect of COVID-19 on influenza vaccination this winter, opinion variables were entered. The three logistic regressions were statistically significant (p < 0.05) and explained 18.3%, 23.5% and 5.7% (Nagelkerke R^2^) of the variance in influenza vaccination, respectively.

Disagreeing to or being neutral about the statement “the influenza vaccine will decrease influenza hospitalization, and spare hospital beds for urgent COVID-19 cases” was the only inverse correlate shared between the 3 groups.

Besides, being afraid of catching the influenza and the COVID-19 together and believing that the influenza vaccine will be effective against COVID-19 significantly influenced the willingness of those who never vaccinated prior to the COVID-19 to vaccinate this winter.

### 7. COVID-19 vaccination

Figure 5A and 5B depict the correlations between general characteristics, KAP regarding COVID-19, and participants’ views, on one side, and COVID-19 vaccination (after the vaccine is endorsed in Lebanon) on the other side.

**Figure 5.**
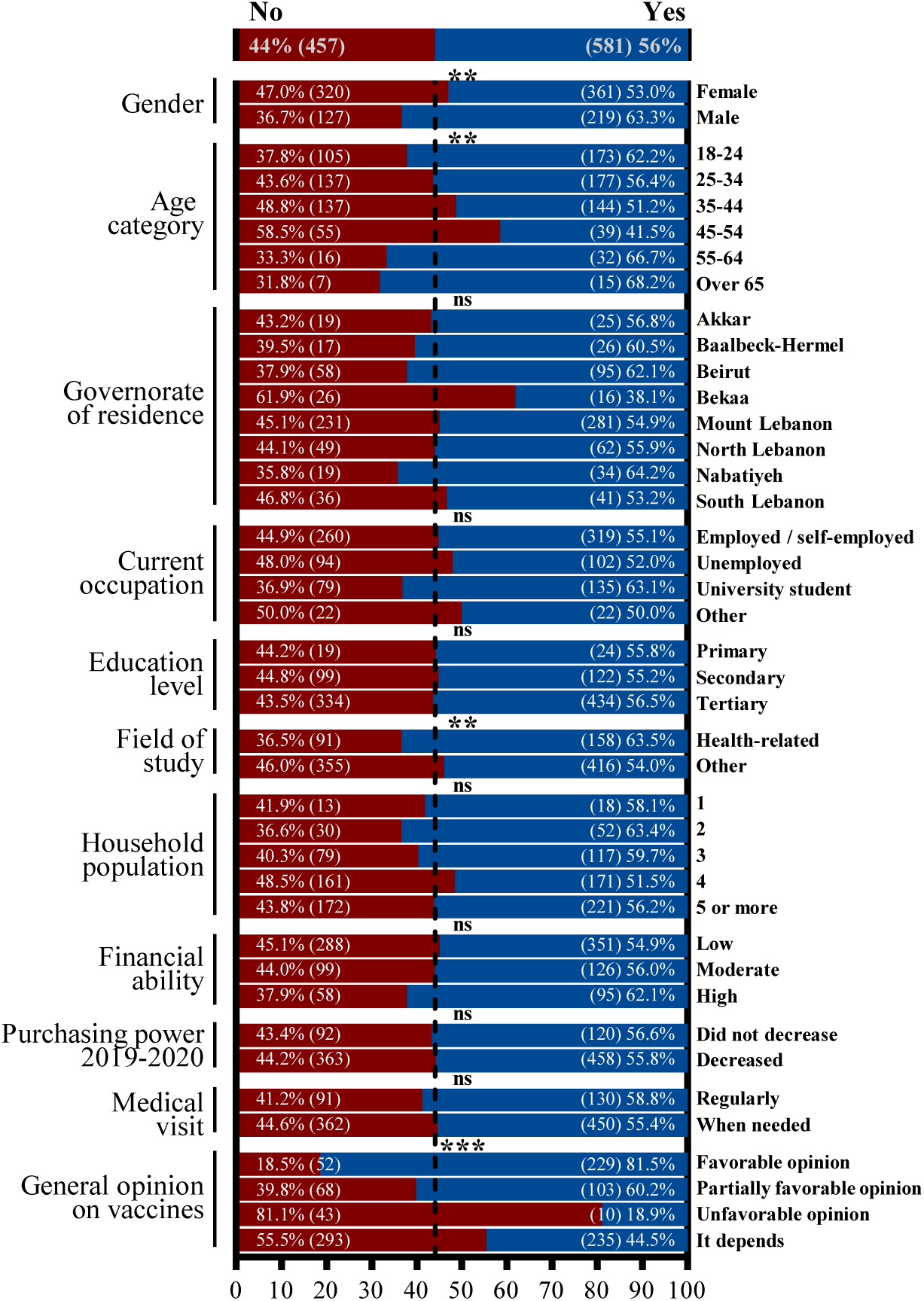
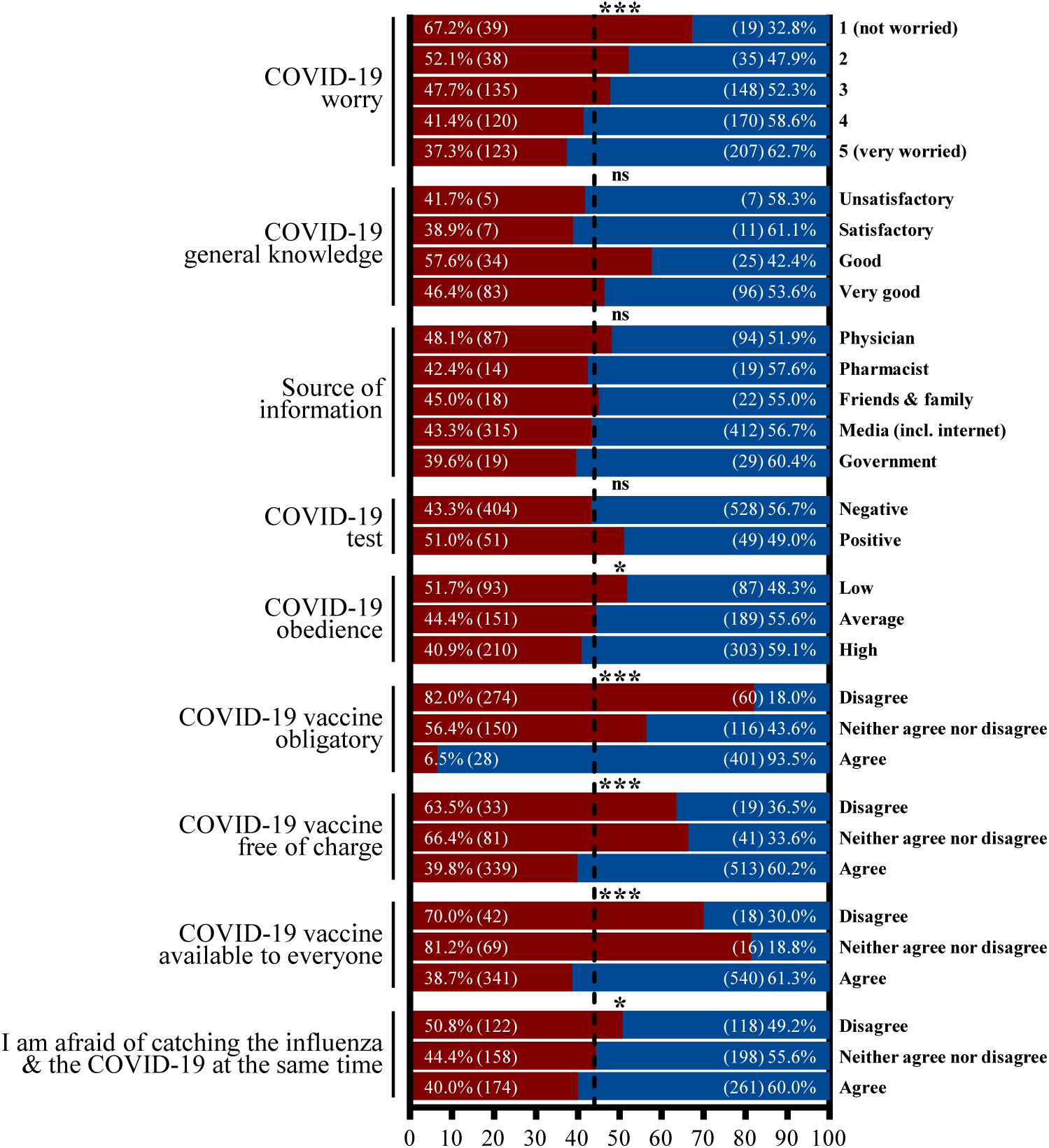
Associations with COVID-19 vaccination. Remark: “COVID-19 obedience” variable (see note 3)

It is obvious that males, old age groups, and individuals with a health-related background are considerably more willing to vaccinate.

Individuals agreeing that the COVID-19 vaccine should be obligatory, are extremely more willing to vaccinate (93.5%) *vs* only 18% and 43.6% of those disagreeing or having no opinion (p<0.001). Comparably, individuals agreeing that they are afraid of catching the influenza and the COVID-19 at the same time are significantly more willing to be vaccinated (p=0.025).

## IV. Discussion

The emergence and spread of severe acute respiratory syndrome coronavirus 2 (SARS-CoV-2) and the resulting coronavirus disease 2019 (COVID-19) led to a global health crisis. Governments worldwide endorsed quarantine measures to slow the spread, protect the most vulnerable, and cope with the tremendous demand on health care services. To stalk the COVID-19 pandemic, regulatory agencies need to support rapid testing and progression of vaccine candidates while in parallel efforts should be directed towards identifying and understanding COVID-19 vaccine hesitancy within distinct populations. Accordingly, we conducted a quantitative cross-sectional study by designing and disseminating a 30-item structured questionnaire. Our study aimed at determining whether the COVID-19 pandemic affected influenza vaccination rate in the general Lebanese adult population with the ultimate goal being to understand the factors behind vaccine hesitancy to shape public health campaigns that could help in increasing the uptake of the influenza and COVID-19 vaccines. To our knowledge, this is the largest study of vaccine confidence in Lebanon, with cross-country comparisons that helped elucidate the public’s vaccination willingness, investigate vaccination confidence, and determine factors behind hesitancy. The sample included 1055 participants with 66.2% being females, and the prevalent age group being 25-34 years old (30.2%). Our results revealed that 5.1% of the participants were anti-vaxxers while the majority (51.3%) could be considered as being hesitant and their willingness was linked to the vaccine and/or production company and the disease. A recent study conducted in Ireland and UK revealed that vaccine resistance/hesitance was evident in 35% and 31% of the respondents, respectively (Murphy et al. 2021). It is noteworthy that prior to the COVID-19 pandemic almost 70% of our study participants never received the flu vaccine within the last five years, with a significant increase in the percentage of participants intending to vaccinate this winter and which our results showed that it was directly linked to the fear of getting a serious COVID-19 infection (results not shown). Perceived risk was previously linked to a change in vaccine hesitancy with at least 30% in proportion willing to vaccinate as risk of infection increases (Baumgaertner et al. 2020).

On the other hand, the education level, the field of study, frequency of medical visits, and attitudes towards vaccination were among the factors in this study with significant impact on the tendency to vaccinate. Tendency to vaccinate was significantly lower in old age brackets compared to the younger participants (p < 0.001), and those having tertiary education representing the dominant within the pro-vaccine group. This was in contrast to the results reported by Baumgaertner et al. (2020) with the older populations being more willing to vaccinate than younger.

There are many other causes given for not getting vaccinated by adults and that are linked to ignorance of the true facts, lack of support and follow up from the healthcare system or baseless fears (Baeyens 2014). Our results came to confirm the aforementioned reasons, where the more the participants were worried about an influenza infection, the more familiar they were with the vaccine and consequently the higher was their tendency to vaccinate. Additionally, suboptimal vaccination was previously linked to affordability and the lack of funding for adult vaccines (Swanson et al. 2015). This agreed with our results, with participants living in households where the purchasing power decreased between 2019 and 2020 were 30% less willing to vaccinate. As the combination of flu and COVID-19 could further overwhelm healthcare settings this winter, there is an urgent need to reform and strengthen public health systems in Lebanon to maximize vaccine availability and affordability and be free and available to the rich and poor alike.

Studying the correlations between knowledge, attitude, and practices regarding COVID-19, and participants’ tendency to vaccinate after introducing COVID-19 vaccine in Lebanon, revealed that males, old age, and having a health-related background demarked those that were more willing to vaccinate. Sex, age, and income level was according to Murphy et al. (2021) the demographic factors that were most significantly associated with vaccine hesitance or resistance. Moreover, 70% of the study participants never received the influenza vaccine within the last 5 years. We think, and according to the received responses, that this hesitancy and resistance could be correlated to knowledge gap and low perceived importance on the impact vaccination that goes beyond the vaccinated individual.

All the aforementioned factors affecting the population’s decision to vaccinate culminated in a worrisome sub-optimal influenza and COVID-19 vaccination rates. Continuing with such rates over the next years will result in a higher burden and mortalities caused by infectious agents against which we have a very affordable and effective prevention measure – vaccination. Combatting the “ideology” of vaccine hesitancy starts with campaigns that stress the success of vaccines on a societal rather than on an individualistic level, highlighting the success of vaccination in eradicating smallpox. We need to promote the integration of health education within all scholastic programs, use technology and media platforms to fight vaccine hesitancy.

### Limitations of the study

The present study has a few limitations. Since the survey was propagated solely in an electronic version, the elderly in our sample were less represented than younger age groups. Moreover, participants were required to self-report their previous vaccination records thus we were unable to double check their accuracy. These rates might not accurately translate in reality for this winter because of the possible lack of influenza vaccines in Lebanon or simply because the participants changed their mind after completing the survey. In spite of all these mentioned limitations, our sample included a considerable number of individuals, of different adult ages, with different educational backgrounds, and most importantly from all 8 governorates in Lebanon to represent as close as possible the general population.

## V. Conclusion

To our knowledge, this study is the largest to evaluate vaccination willingness among the general adult population in Lebanon emphasizing on influenza and endorsing COVID-19 programs. We observed a significant increase in the predicted tendency to be vaccinated against influenza for this winter relative to the rate prior to the COVID-19 pandemic but is still alarmingly sub-optimal. We believe that maximizing the availability and affordability of vaccines, providing public educational campaigns, involving healthcare workers to educate the public, and implementing immunization reminder-recall systems are much needed in the time of the COVID-19 pandemic.

## Supporting information

Supplemental Survey

## Data Availability

All the data is available and included

## Notes

1. Participants were asked about their family income as well as the number of people living in their household. Consequently, the family incomes were divided by the number of people living in the household to determine the income per person per month in Lebanese pounds (LBP) which was categorized into either low (less than 1 000 000 LBP), moderate (between 1 000 000 and 3 000 000 LBP), or high financial standing (above 3 000 000 LBP).
2. Ten questions regarding participants’ general knowledge about influenza and COVID-19 were scored either 0 for a wrong answer or 1 for a correct answer. The scores were added up to 5 possible points for each disease which were consequently grouped into 4 categories: unsatisfactory (0/5, 1/5, or 2/5), satisfactory (3/5), good (4/5), and very good (5/5).
3. Participants’ obedience during the COVID-19 pandemic was self-reported on a 5-point scale for 4 phases of the pandemic: Phase I (In March 2020: Beginning of COVID-19 cases in Lebanon), Phase II (From April to May 2020: Rise in the COVID-19 cases in Lebanon), Phase III (From May to August 2020: Drop in the COVID-19 cases in Lebanon), and Phase IV (After August 2020 [Beirut blast]: Rise in the COVID-19 cases in Lebanon). The scores were then averaged for each individual for analytic purposes and classified into low (2.5/5 and below), average (between 2.5/4 and 4/5), high (4/5 and above).

## Notes

### Competing Interest Statement

The authors have declared no competing interest.

### Clinical Trial

It's a survey not a clinical trial

### Funding Statement

Not applicable

### Author Declarations

Institutional Review Board (IRB) at the Lebanese American University (approval number LAU.SAS.ST3.4/Nov/2020).

## References

Baeyens, J-P. (2014). Ensuring the willingness to vaccinate and be vaccinated. Expert Review of Vaccines 9:sup3, 11–14.

Baumgaertner, B., Ridenhour, B.J., Justwan, F., Carlisle, J.E., & Miller, C.R. (2020). Risk of disease and willingness to vaccinate in the United States: A population-based survey. PLOS Medicine 17(10): e1003354.

Centers for Disease Control and Prevention. (2018). Estimates of Influenza Vaccination Coverage among Adults—United States, 2017–18 Flu Season. Retrieved February 5, 2021

Central Administration of Statistics. (2019). Population Statistics. http://www.cas.gov.lb/index.php/demographic-and-social-en/population-en. Retrieved February 5, 2021

El Khoury, G. & Salameh, P. (2015). Influenza Vaccination: A Cross-Sectional Survey of Knowledge, Attitude and Practices among the Lebanese Adult Population. International Journal of Environmental Research and Public Health 12(12):15486–97.

European Centre for Disease Prevention and Control. (2017). Seasonal influenza vaccination in Europe – Vaccination recommendations and coverage rates for eight influenza seasons (2007–2008 to 2014–2015). Retrieved February 5, 2021

Coronaviridae Study Group of the International Committee on Taxonomy of Viruses., Gorbalenya, A.E., Baker, S.C. et al. (2020). The species Severe acute respiratory syndrome-related coronavirus: classifying 2019-nCoV and naming it SARS-CoV-2. Nature Microbiology 5(536–544).

Ministry of Public Health. (2021). Novel Coronavirus 2019: https://www.moph.gov.lb/en/Pages/2/24870/novel-coronavirus-2019. Retrieved February 5, 2021

Ministry of Public Health. (2021). Lebanon National Drugs Database: https://www.moph.gov.lb/en/Drugs/index/3/4848/lebanon-national-drugs-database. Retrieved February 5, 2021

Murphy, J., Vallières, F., Bentall, R.P., Shevlin, M., McBride, O., Hartman, T.K., McKay, R., Bennett, K., Mason, L., Gibson-Miller, J., Levita, L., Martinez, A.P., Stocks, T.V.A., Karatzias, T., Hyland, P. (2021). Psychological characteristics associated with COVID-19 vaccine hesitancy and resistance in Ireland and the United Kingdom. Nature Communications 12(29).

Nafziger, A.N., & Pratt, D.S. (2014). Seasonal influenza vaccination and technologies. 54(7):719–31.

Solomon, D.A., Sherman, A.C., & Kanjilal, S. (2020). Influenza in the COVID-19 Era. JAMA 324(13):1342–1343.

Swanson, K.A., Schmitt, J.H., Jansen, K.U., and Anderson, A.S. (2015). Adult vaccination. Human Vaccines & Immunotherapeutics 11(1):150–5.

U.S. Food & Drug Administration. (2021). COVID-19 Vaccines: https://www.fda.gov/emergency-preparedness-and-response/coronavirus-disease-2019-covid-19/covid-19-vaccines#news. Retrieved February 5, 2021

World Economic Forum. (2020). Three in Four Adults Globally Say They’d Get a Vaccine for COVID-19 – But Is This Enough? Retrieved February 5, 2021

World Health Organization. (2018). Ask the expert: Influenza Q&A: https://www.who.int/en/news-room/fact-sheets/detail/influenza-(seasonal). Retrieved February 5, 2021

World Health Organization. (2021). WHO Coronavirus Disease (COVID-19) Dashboard: https://covid19.who.int/?gclid=CjwKCAiA9bmABhBbEiwASb35VyTkisNf3cEjY2SmOJvPmOoMse2VWYylOHnDKrvPB9xVYNjtdKBfVRoCt7cQAvD_BwE. Retrieved February 5, 2021

