## Supplemental Figures for "COVID-19 and Influenza: Vaccination Before and During the Pandemic among the Lebanese Adult Population"

**Appendices**

Appendix A. Supplementary Figures

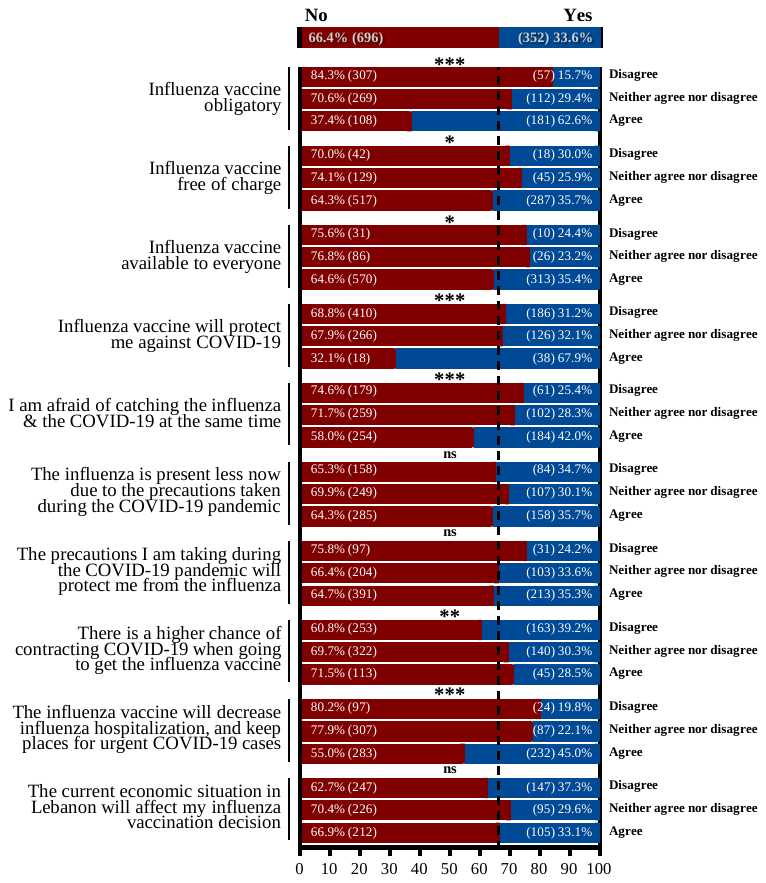

**Supplementary Figure 1.** Participants’ views and influenza vaccination during the COVID-19 pandemic**.**

Appendix B. Call for action to minimize vaccine hesitancy.

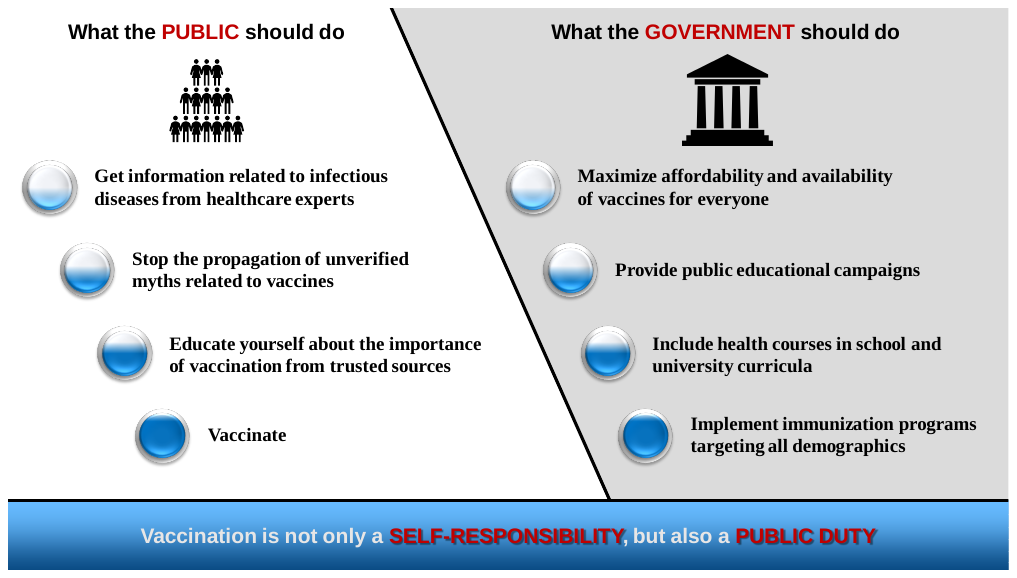
