## Supplemental Survey for "COVID-19 and Influenza: Vaccination Before and During the Pandemic among the Lebanese Adult Population"

### FLUVACO – Survey Questions

#### Section A: General information

1. What is your gender
   - Male
   - Female

2. How old are you?
   - Under 18
   - 18-24
   - 25-34
   - 35-44
   - 45-54
   - 55-64
   - 65+

3. What is your area (Mouhafaza) of residency?
   - Akkar
   - Baalbeck-Hermel
   - Beirut
   - Bekaa
   - Mount Lebanon
   - North Lebanon
   - Nabatiyeh
   - South Lebanon

4. Which of the following best describes your current occupation?
   - Employed
   - Voluntarily unemployed
   - Involuntarily unemployed
   - Volunteer
   - School student
   - University student
   - Retired

5. What is your highest education level ?
   - Uneducated
   - Elementary school
   - Middle school (Brevet)
   - Technical baccalaureate (BT-TS)
   - High school (Baccalaureate)
   - Bachelor or equivalent
   - Master or equivalent
   - Doctorate or equivalent

6. What is / was your field of studies?
   - Healthcare / Medicine / Pharmacy / Life Sciences
   - Other

7. How many people are currently living in your household, including yourself?
   - 1
   - 2
   - 3
   - 4
   - 5 or more

FLUVACO

8. What is your total combined household monthly income? If you do not know your exact income, please estimate
   - No income
   - Less than 1,000,000 LBP
   - Between 1,000,000 and 2,500,000 LBP
   - Between 2,500,000 and 5,000,000 LBP
   - Between 5,000,000 and 7,500,000 LBP
   - Between 7,500,000 and 10,000,000 LBP
   - More than 10,000,000 LBP

9. What happened to the purchasing power of your household since the start of the economic crisis in Lebanon (around September 2019)?
   - Increased
   - Decreased
   - Neither increased nor decreased

10. Do you have any of the following chronic disease conditions?

| | Yes | No |
|---|---|---|
| Diabetes mellitus | | |
| Cancer | | |
| Respiratory disease | | |
| Kidney disease | | |
| Cardiovascular disease | | |
| Liver disease | | |
| Immunodeficiency disorder | | |

11. How frequently do you visit a physician?
    - Routinely
    - Once per 6 months
    - Once per year
    - When needed

12. How do you feel about vaccines in general?
    - Favorable opinion regarding any vaccine (Pro-vaxxer)
    - Partially favorable opinion regarding any vaccine
    - Unfavorable opinion regarding any vaccine (anti-vaxxer)
    - It depends on the disease / vaccine / production company

FLU VAC

#### Section B: Influenza and Influenza vaccine

1. General knowledge about influenza (seasonal flu)

| | Yes | No | Not sure |
|---|---|---|---|
| It is caused by a virus | | | |
| It can spread from person to person | | | |
| It can be prevented | | | |
| It can cause fever | | | |
| It can change / alter from year to year | | | |

2. How do you feel about an influenza infection ?

Not worried ○ 1 ○ 2 ○ 3 ○ 4 ○ 5 Very worried

3. In the last five years, how many times did you get the influenza vaccine?
   - ○ Every year (then questions 4, 6 of section B)
   - ○ Sometimes (then questions 4,5, and 6 of section B)
   - ○ Never (then question 5 of section B)

4. What is the main reason why you get the influenza vaccine?
   - ○ I want to protect myself, my friends, and my family
   - ○ I want to protect the society
   - ○ The vaccine is obligatory in my work/college/school
   - ○ My physician advised me to get it

5. What is the main reason that you do not get the vaccine?
   - ○ I am healthy
   - ○ It is expensive
   - ○ I am afraid of the side effects
   - ○ I got an influenza shot once, so I do not need to get it again

6. When do you usually get the influenza vaccine?
   - ○ Before the influenza season
   - ○ During the influenza season
   - ○ After the influenza season
   - ○ When sick with the influenza
   - ○ After getting sick with the influenza

7. How often do you take the following precautions (other than vaccination) to protect yourself from the influenza?

| | Never | Rarely | Sometimes | Often | Always |
|---|---|---|---|---|---|
| Disinfecting my hands and face | | | | | |
| Keeping distance from someone showing symptoms | | | | | |
| Quarantining during the influenza season (mainly winter) | | | | | |
| Taking medications and supplements to boost my immune system | | | | | |

8. How familiar are you with the influenza vaccine (how it works, efficacy, etc.)?

Not familiar at all ○ 1 ○ 2 ○ 3 ○ 4 ○ 5 Very familiar

FLUVACO

9. What is your main source of information regarding the influenza vaccine?
    - Physician
    - Pharmacist
    - Friends and family
    - Media (including the internet)
    - Government

FLUVACO

#### Section C: COVID-19

1. General knowledge about COVID-19 (Corona)

| | Yes | No | Not sure |
|---|---|---|---|
| It is caused by a virus | | | |
| It can spread from person to person | | | |
| It can be prevented | | | |
| It is the same as the influenza | | | |
| It can change / alter in the future | | | |

2. How do you rate your obedience (out of 5) in taking precautions regarding the COVID-19 pandemic?

| | 1 (not good) | 2 | 3 | 4 | 5 (very good) |
|---|---|---|---|---|---|
| In March 2020 (Phase I: beginning of the cases in Lebanon) | | | | | |
| From April 2020 to May 2020 (Phase II: rise in the cases in Lebanon) | | | | | |
| From May 2020 to August 2020 (Phase III: drop in the cases in Lebanon) | | | | | |
| From August 2020 till present (Current phase: after the Beirut explosion) | | | | | |

3. Up to this day, were you tested positive for COVID-19 through a PCR?
   - Yes
   - No

4. How do you feel about a COVID-19 infection ?

   Not worried ○ 1 ○ 2 ○ 3 ○ 4 ○ 5 Very worried

5. What is your main source of information regarding COVID-19?
   - Physician
   - Pharmacist
   - Friends and family
   - Media (including the internet)
   - Government

FLUVACO

#### Section D: Influenza – COVID-19 intersection

1. What will you do this winter regarding influenza vaccination ?
   - Surely get the vaccine
   - Most probably get the vaccine
   - Most probably not get the vaccine
   - Not get the vaccine

2. What do you think about the following?

| | Disagree | Neither agree nor disagree | Agree |
|---|---|---|---|
| The influenza vaccine will protect me from COVID-19 | | | |
| I am afraid of catching the influenza and the COVID-19 at the same time | | | |
| The influenza is present less now due to the precautions during the COVID-19 pandemic | | | |
| The precautions I am already taking during the COVID-19 pandemic will also protect me from the influenza | | | |
| There is a higher chance of contracting COVID-19 when going to get the influenza vaccine | | | |
| The influenza vaccine will decrease influenza hospitalization, and keep places for urgent COVID-19 cases | | | |
| The current financial crisis in Lebanon will affect my influenza vaccination decision | | | |

3. To answer the following questions, suppose that a vaccine to prevent a COVID-19 infection is produced and approved for usage

   Once the COVID-19 vaccine is available, I will …
   - get both the influenza and the COVID-19 vaccines
   - get the influenza vaccine only
   - get the COVID-19 vaccine only
   - not get the influenza vaccine nor the COVID-19 vaccine

4. How do you feel about the following for this winter ?

| | Disagree | Neither agree nor disagree | Agree |
|---|---|---|---|
| The influenza vaccine should be obligatory | | | |
| The COVID-19 vaccine should be obligatory | | | |
| The influenza vaccine should be free of charge | | | |
| The COVID-19 vaccine should be free of charge | | | |
| The influenza vaccine should be available for everyone | | | |
| The COVID-19 vaccine should be available for everyone | | | |

FLUVACO
